# Heparin-induced thrombocytopenia is associated with a high risk of mortality in critical COVID-19 patients receiving heparin-involved treatment

**DOI:** 10.1101/2020.04.23.20076851

**Authors:** Xuan Liu, Xiaopeng Zhang, Yongjiu Xiao, Ting Gao, Guangfei Wang, Zhongyi Wang, Zhang Zhang, Yong Hu, Qincai Dong, Songtao Zhao, Li Yu, Shuwei Zhang, Hongzhen Li, Kaitong Li, Wei Chen, Xiuwu Bian, Qing Mao, Cheng Cao

## Abstract

**Background:** Coronavirus infectious disease 2019 (COVID-19) has developed into a global pandemic. It is essential to investigate the clinical characteristics of COVID-19 and uncover potential risk factors for severe disease to reduce the overall mortality rate of COVID-19.

**Methods:** Sixty-one critical COVID-19 patients admitted to the intensive care unit (ICU) and 93 severe non-ICU patients at Huoshenshan Hospital (Wuhan, China) were included in this study. Medical records, including demographic, platelet counts, heparin-involved treatments, heparin-induced thrombocytopenia-(HIT) related laboratory tests, and fatal outcomes of COVID-19 patients were analyzed and compared between survivors and nonsurvivors.

**Findings:** Sixty-one critical COVID-19 patients treated in ICU included 15 survivors and 46 nonsurvivors. Forty-one percent of them (25/61) had severe thrombocytopenia, with a platelet count (PLT) less than 50×10^9^/L, of whom 76% (19/25) had a platelet decrease of >50% compared to baseline; 96% of these patients (24/25) had a fatal outcome. Among the 46 nonsurvivors, 52·2% (24/46) had severe thrombocytopenia, compared to 6·7% (1/15) among survivors. Moreover, continuous renal replacement therapy (CRRT) could induce a significant decrease in PLT in 81·3% of critical CRRT patients (13/16), resulting in a fatal outcome. In addition, a high level of anti-heparin-PF4 antibodies, a marker of HIT, was observed in most ICU patients. Surprisingly, HIT occurred not only in patients with heparin exposure, such as CRRT, but also in heparin-naïve patients, suggesting that spontaneous HIT may occur in COVID-19.

**Interpretation:** Anti-heparin-PF4 antibodies are induced in critical COVID-19 patients, resulting in a progressive platelet decrease. Exposure to a high dose of heparin may trigger further severe thrombocytopenia with a fatal outcome. An alternative anticoagulant other than heparin should be used to treat COVID-19 patients in critical condition.

**Funding:** This investigation was supported by grants 2016CB02400 and 2017YFC1201103 from the National Major Research and Development Program of China.

## Introduction

Coronavirus infectious disease 2019 (COVID-19), caused by a novel coronavirus (SARS-CoV-2) that is structurally related to the virus that causes severe acute respiratory syndrome (SARS), has developed into a global pandemic, with two million infections and 130000 deaths.^1-3^ Currently, the mortality of COVID-19 has reached as high as 13%, such as in France and Italy.^4^ In China, approximately 81% of COVID-19 patients have been mild cases, whereas 14% have been severe cases with acute respiratory distress syndrome (ARDS) that require ventilator support, and 5% have been critical patients who need intensive care unit (ICU) admission.^5^ The survival rate of critical patients is no more than 50%. Elderly people and people who have underlying medical conditions are at higher risk for developing more serious complications, such as septic shock, ARDS, acute kidney injury (AKI), and fulminant myocarditis.^6,7^ Approximately 81% of deaths occurred among people aged >60 years in China.^5^

As a new infectious disease, COVID-19 is characterized by severe acute respiratory injury and hyperinflammation-related syndromes. The poor prognosis predictors uncovered by retrospective studies include hypercytokinemia^6^, significantly elevated hallmarks of an inflammatory response (e.g. IL-6, C-reactive protein (CRP), and serum ferrin) and organ failure (e.g., aspartate or alanine transaminase (AST or ALT), cardiac troponin, and blood urea nitrogen (BUN)),^7,8^ and SARS-CoV-2-related RNAemia.^9^ Additionally, severe thrombocytopenia, elevated neutrophils, and reduced lymphocyte have also been reported extensively in critical patients.^10,11^ Combined with progressive pulmonary lesions on CT imaging, these findings may be helpful to establish more appropriate clinical guidance for critically ill cases.

A low platelet count or thrombocytopenia is associated with an increased risk of severe disease and mortality in patients with SARS and COVID-19 for reasons that are not fully understood.^12^ Heparin-induced thrombocytopenia (HIT) is a well-recognized complication of heparin therapy, and can occur spontaneously, independent of heparin.^13-15^ HIT is caused by the binding of heparin to platelet factor 4 (PF4) released from activated platelets, as antibodies against the heparin-PF4 complex are induced in some patients. Subsequently, the antibody-PF4-heparin complex binds to the platelet FcγIIa receptor, inducing platelet activation and aggregation, activation of the coagulation pathways and an eventual loss of circulating platelets, with a prothrombotic state.^16,17^ Nevertheless, HIT can be induced in the absence of proximate heparin exposure, so-called spontaneous HIT, by negatively charged bacterial, nucleic acids, and hypersulfated chondroitin.^18-20^ Most patients who develop spontaneous HIT have proximate episodes of infection or major surgery.^18^

In this study, 61 critical ICU patients and 93 severe non-ICU patients hospitalized from February 8 to April 03 at Huoshenshan Hospital (Wuhan, China), an emergency hospital established in for COVID-19 patients, were included. We found that a substantial percentage of the critical ICU patients (31·1%, 19/61) who showed severe, progressive decreases in platelet counts, often with fatal outcomes. Further analysis showed an elevated level of anti-heparin-PF4 antibodies (HIT antibodies) in critical patients, which suggested that HIT contribute greatly to the fatal outcome in COVID-19 patients in critical condition.

## Methods

### Patients and data collection

Sixty-one patients with confirmed COVID-19 in critical condition were admitted to Wuhan Huoshengshan Hospital, a hospital established for severe and critical COVID-19 patients, from February 8 to March 18. Every patient had stayed in the intensive care unit for more than three days (herein and after referred to as ICU patients) and had at least three consecutively detected platelet counts data. Another 93 patients with severe COVID-19 who had never stayed in the ICU were randomly selected from the hospital (herein and after referred to as non-ICU patients). The diagnosis of COVID-19 was made according to Chinese Clinical Guidance for COVID-19.^21^ Viral RNA was confirmed in all patients by reverse transcription real-time PCR in the clinical laboratory of the hospital, except for patient P11 who was RNA-positive before admission to the hospital but turned to be negative during the hospitalization. The clinical outcomes were recorded up to April 03, 2020. Routine blood tests and coagulation tests were performed in the hospital laboratory on admission and during the hospitalization. Demographic data for the patients, including information on age and gender, are shown in supplementary table S1. This study was approved by the Ethics Committee of Huoshenshan Hospital (Wuhan, China).

### Anti-heparin-PF4 antibody assay (HIT assay)

Human anti-heparin-PF4 complex antibodies (IgG) were detected using an antigen-sandwiched enzyme-linked immunosorbent assay (ELISA) kit produced by Jonln Biological Company (Cat JL12174). Briefly, 50 μl of serum was added to a microwell coated with heparin-PF4 complex, incubated at 37°C and washed with washing buffer. Then, horseradish peroxidase (HRP)-conjugated hepairn-PF4 was added, incubated, and carefully washed, HRP activity was determined by the ability to convert the substrate 3,3′,5,5′-tetramethylbenzidine (TMB) to a blue products. The amount of HIT antibodies was quantified using a set of calibration standards provided by the manufacturer.

### Serum C3a determination

Serum C3a levels were determined by a two-antibody sandwich ELISA similar to the HIT assay. In the assay, the microplate well was coated with anti-C3a antibody (capturing antibody), and the presence of C3a in patient’s serum was quantified with an HRP-conjugated C3a antibody.

### Statistical analysis

For the survival rate and qualitative data comparisons among different groups, the *P-*value was calculated using the Chi-Square Test (χ^2^) or Fisher’s exact test in R (version 3.6.2). A *t*-test was used to determine the significant difference. *P*<0.05 was considered statistically significant. Data were presented as mean ± SD or mean ± SEM.

## Results

### Severe, progressive thrombocytopenia was observed in critical COVID-19 patients

To address the fact that many patients in the ICU received transfusions, the complete blood count data during hospitalization were analyzed. As expected, decreased red blood cell count (RBC) and hemoglobin (RGB) were observed in most ICU patients (figure 1A). To our surprise, a progressive, severe decrease in platelet count (PLT) occurred in critical ICU patients with fatal outcomes (figure 1A and S1A, and table 1), but rarely occurred in ICU survivors (figure 2 and table 1) and non-ICU patients with severe COVID-19 (figure S1B). As shown in table 1, severe thrombocytopenia (PLT≤50×10^2^/L) was observed in 1·1% of non-ICU patients (1/93), whereas it occurred in 41% of ICU patients (25/61), of whom 96% were ICU nonsurvivors (24/25). Moreover, approximately 52·2% of the ICU nonsurvivors (24/46) had a PLT count less than 50×10^2^/L, compared to 6·7% among ICU survivors (1/15) (table 1). Notably, 21·7% of ICU nonsurvivors (10/46) had critical thrombocytopenia, with PLT less than 10×10^2^/L in the very late phase (1-3 days before death) of the disease (figure 1B and table 1). Significant PLT decreases (a decrease of >50% at the last sampling date compared to that at the admission date) were also observed in 56·5% of ICU nonsurvivors (26/46) but in neither ICU survivors nor non-ICU patients (figure 1C).

**Table 1.**
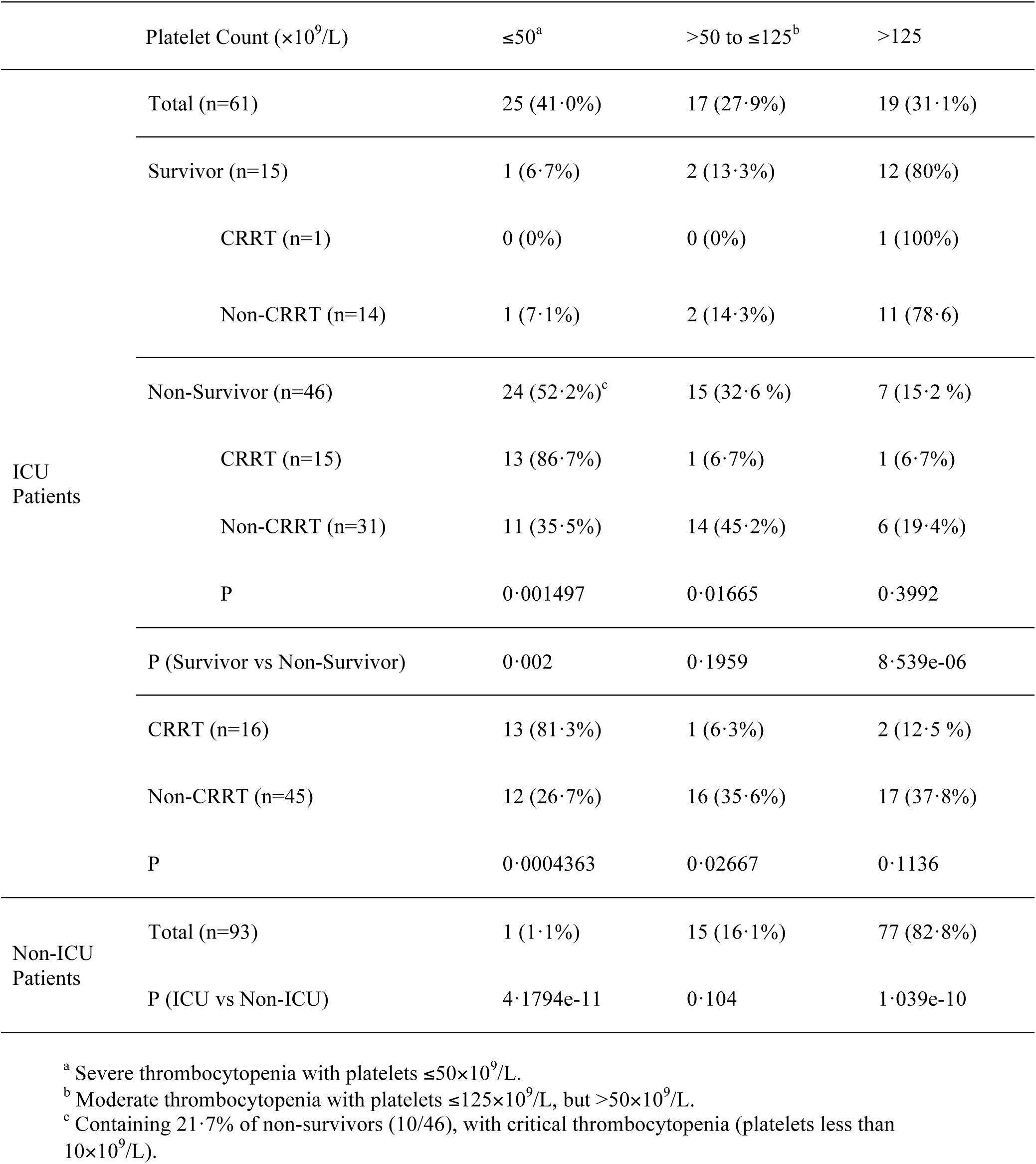
Platelet counts, CRRT treatments, and outcomes.

**Figure 1.**
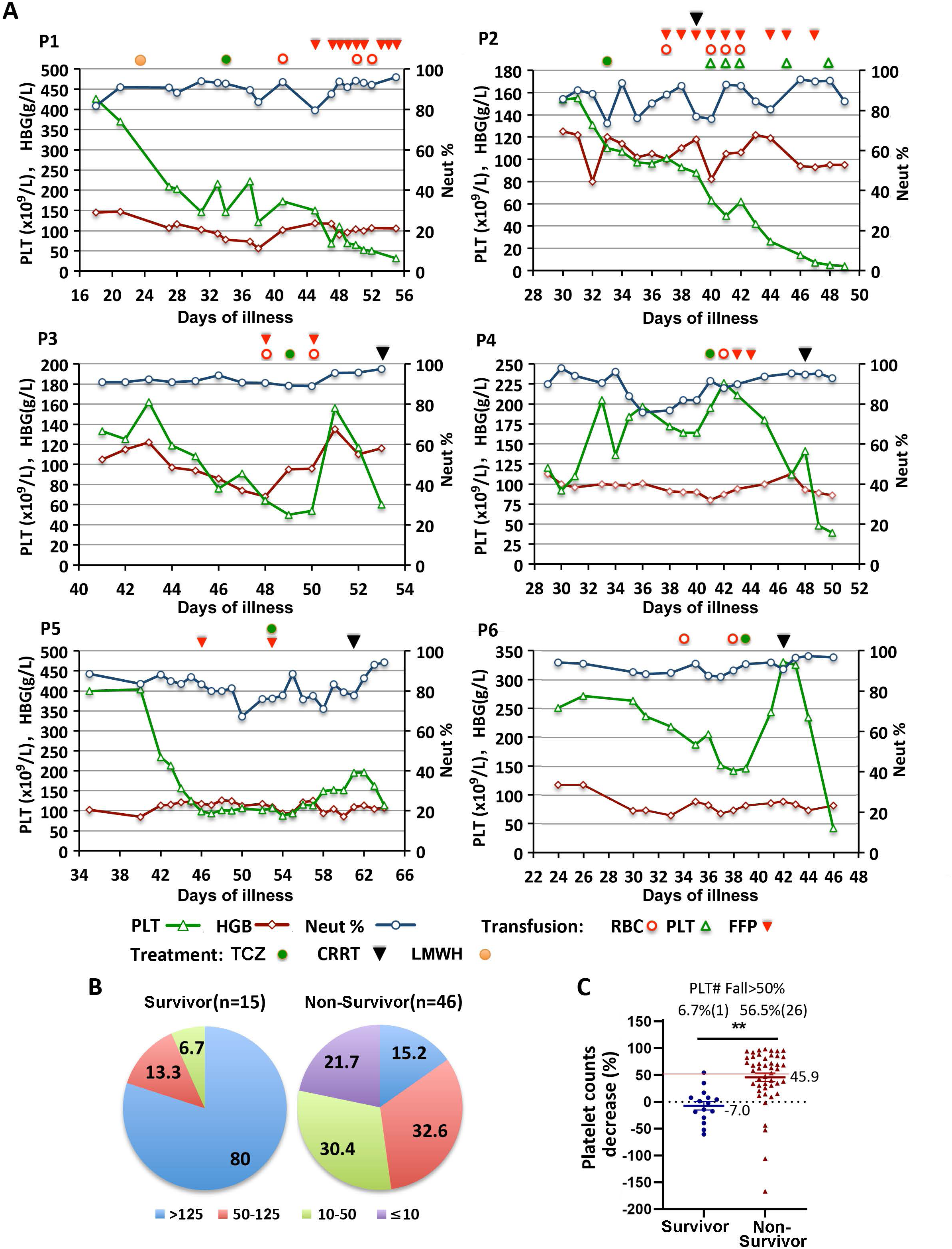
Severe, progressive thrombocytopenia in nonsurviving COVID-19 patients in the ICU. (A) Day-by-day variations of platelet counts, hemoglobin levels, and neutrophils percentages in six nonsurviving patients (P1-P6) during ICU admission are shown. Treatments with tocilizumab, transfusion, CRRT, and heparin exposure is labeled in the figure. PLT, platelet (×10^2^/L); HGB, hemoglobin (g/L); Neut%, neutrophils percentage; TCZ, tocilizumab treatment; CRRT, continuous renal replacement therapy; LMWH, low-molecular-weight heparin treatment; RBC, red blood cells; FFP, fresh frozen plasma. (B) Platelet count ranges (×10^2^/L) in 15 surviving and 46 nonsurviving COVID-19 patients. The percentage of patients with each platelet counts range is shown in the figure. (C) Platelet count decrease of each patient was calculated by (PLT#(admission day)-PLT#(the last day))/PLT#(admission day)*100%. The percentage of surviving or nonsurviving COVID-19 patients with a >50% decrease of platelet is shown in the figure. The data is shown as mean ± SEM and analyzed using a two-tailed Student’s *t-*test. Differences were considered significant at *\*\*P*<0·01, as indicated.

**Figure 2.**
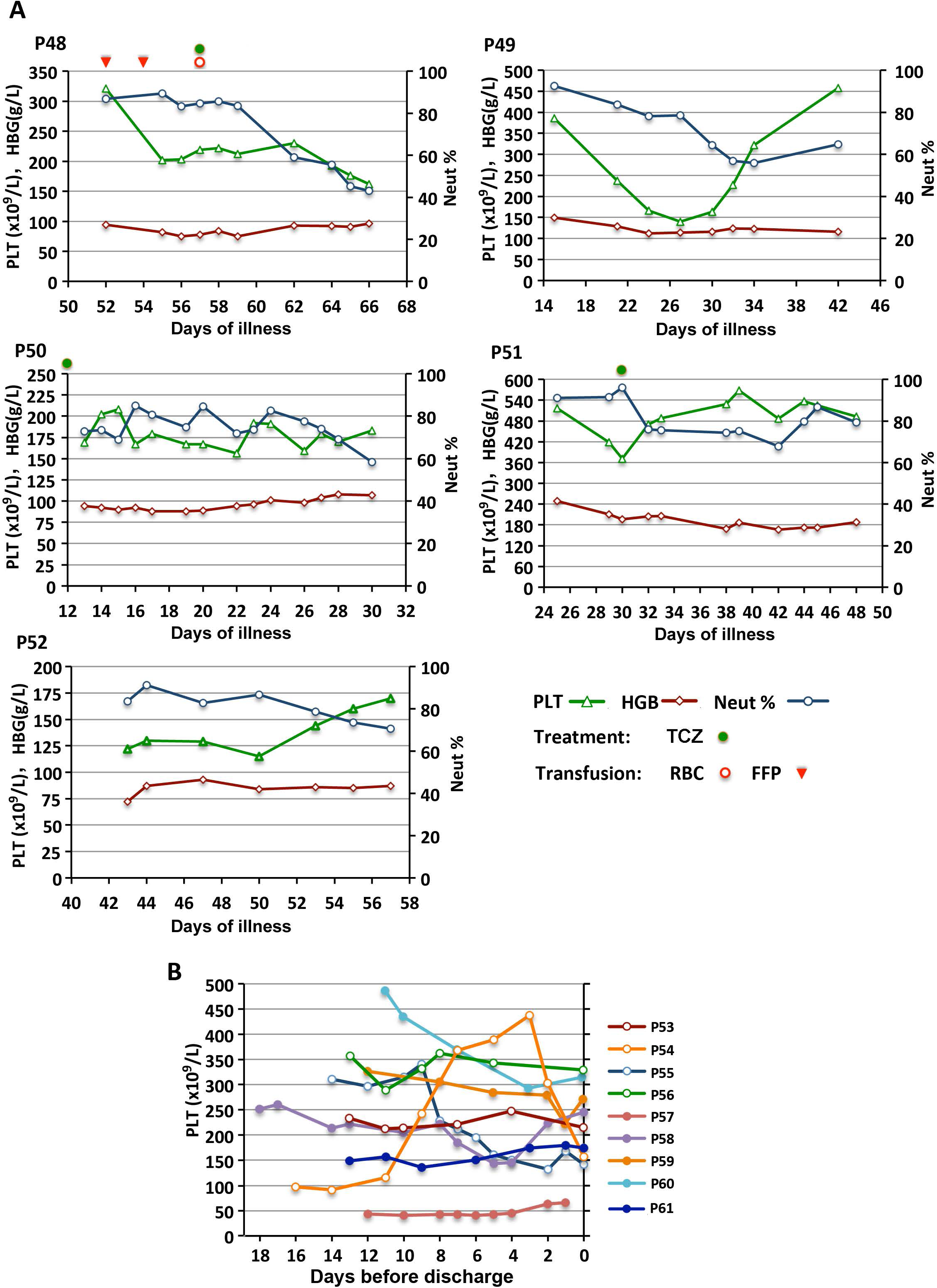
Platelet counts in 14 surviving ICU patients without CRRT treatment. (A) Day-by-day variations of PLT#, HGB, and Neut% in 5 surviving patients without CRRT treatment (P48-P52) during ICU admission is shown in the figure. Treatments are labeled as in figure 1A. (B) The trend lines of day-by-day platelet count in other surviving ICU patients without CRRT treatment (P53-P61).

Analysis the variations in PLT in nine ICU nonsurvivors (patient P1-P6 and P16-P18) during hospitalization in detail found that PLT began to decrease 20-40 days after the onset of the illness and progressively decreased to a critical condition in the next 5-30 days (figure 1A and S1A). Countermeasures such as transfusion with blood cells, platelets or fresh frozen plasma (FFP) showed little if any effect on the PLT decreases (figure 1A and S1A), suggesting that progressive PLT exhaustion occurred in dying COVID-19 patients for unknown reason other than blood coagulation factor exhaustion. Administration of tocilizumab (TCZ), an inhibitor of inflammation that has been suggested for used in COVID-19 by Chinese guidelines for COVID-19, seemed to be able to reverse the changes in PLT transiently in patient P1-P6, except P2, but severe thrombocytopenia recurred a few days after the administration (figure 1A). As a control, only one out of fifteen ICU survivors (patient P57) showed extremely low PLT, but without a progressive PLT decrease (figure 2A and 2B). These results suggested that severe thrombocytopenia with progressively platelet count decrease occurred in most critical COVID-19 patients before a fatal outcome.

### Continuous renal replacement therapy and heparin exposure induce critical thrombocytopenia

Continuous renal replacement therapy (CRRT) is commonly used in the ICU for correction of metabolic acidosis, removal of cytokines and other indications in critical COVID-19 patients. However, among 16 CRRT patients (P2-P17), CRRT triggered a sharp decrease in PLT in most patients (figure 1A, 3A, S1A, and S1C). Notably, among the CRRT-treated COVDI-19 patients, patient P15, the only survivor of CRRT with moderate pneumonia but critical kidney failure, and patient P11, who was viral RNA-negative from the time of admission to the hospital, showed no decline in PLT after CRRT treatment compared with other CRRT treated COVID-19 patients (figure 3A), suggesting that CRRT could induce a significant decrease in PLT in critical patients with severe COVID-19 characteristic pneumonia.

**Figure 3.**
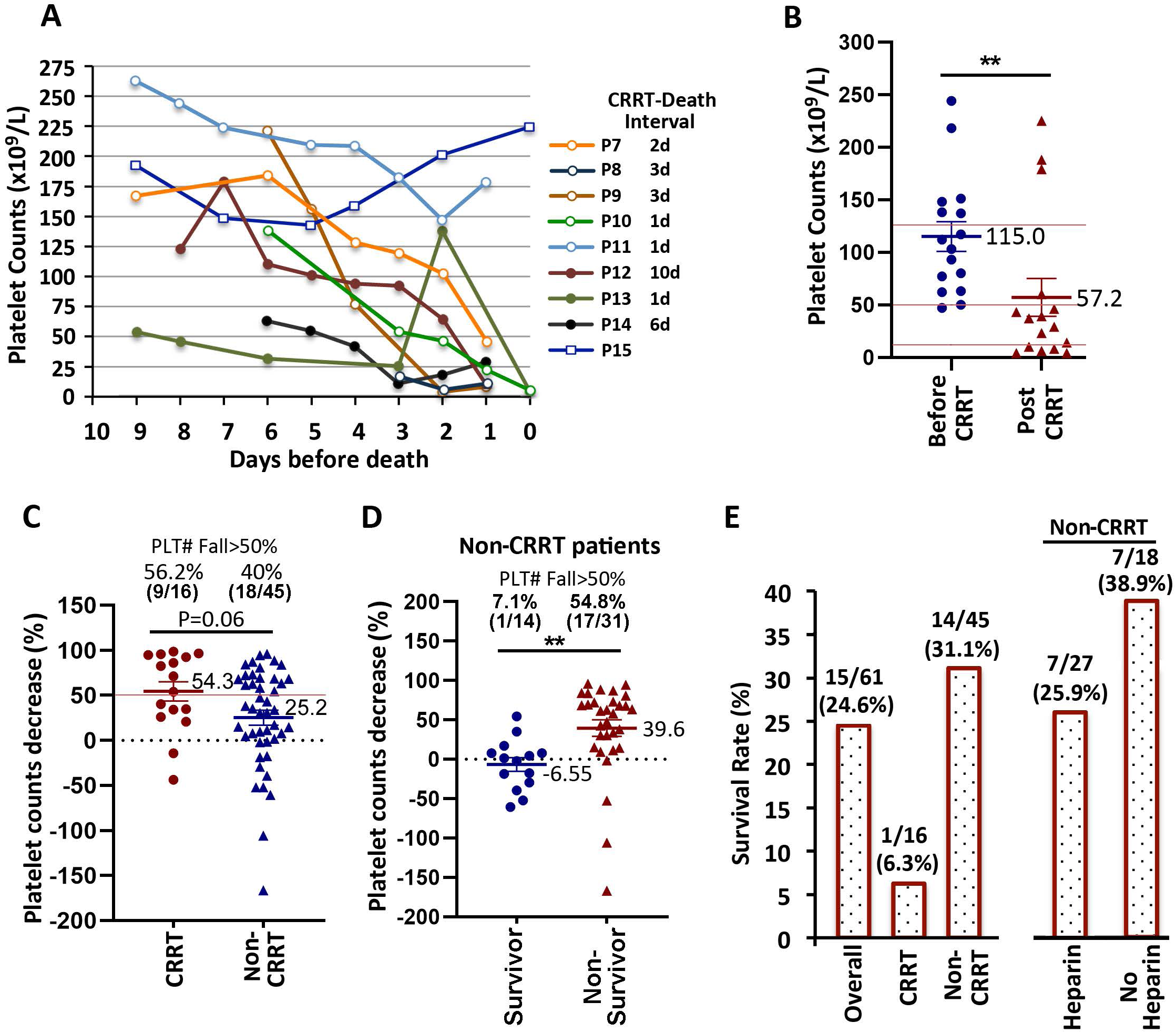
CRRT treatment induces severe thrombocytopenia and death in critical COVID-19 patients. (A) The trend lines of day-by-day platelet count variation in CRRT-treated patients (P7-P15). P15 is the only surviving patient. (B) The average platelet counts before (1 day before or the same day as CRRT) and after (the last available data before death) CRRT. (C) The decrease percentage of platelet count in COVID-19 patients treated with or without CRRT. The average PLT decrease and the percentage of CRRT or non-CRRT treated patients with a >50% decrease of platelet is demonstrated. (D) The decrease percentage of platelet count in recovered (surviving) or dead (nonsurviving) COVID-19 patients without CRRT treatment. The average PLT decrease in surviving or nonsurviving patients is demonstrated. (E) Survival rate in COVID-19 patients treated with or without CRRT or heparin. The data are presented as the mean ± SEM and were analyzed using a two-tailed Student’s *t-*test. Differences were considered significant at *\*\*P*<0·01, as indicated.

Accordingly, a seriously reduced PLT to less than 50×10^2^/L was observed in 81·3% of CRRT patients (13/16) after CRRT therapy, of whom 46·2% (6/13) further decreased to a level less than 10×10^2^/L in a few days after CRRT treatment (figure 3B and S1C). Comparing with that, only 26·7% in non-CRRT ICU patients (12/45), and 1·1% in non-ICU patients (1/93) had a decreased PLT less than 50×10^2^/L (table 1). Moreover, a significant PLT fall of more than 50% was also observed in CRRT patients (56·2%, 9/16) compared to non-CRRT ICU patients (40%, 18/45) (*P*=0·061) (figure 3C), particularly for non-CRRT ICU survivors (7·1%, 1/14) (figure 3D). For non-CRRT patients, the average decrease in PLT in nonsurviving patients was 39·6%, compared to -6·55% (i.e., increased by 6·55%) in survivors (figure 3D), indicating that a significant decrease in PLT may be a risk factor for high mortality in COVID-19.

In concert with severe thrombocytopenia, 11 out of 16 ICU patients treated with CRRT died within three days post CRRT, and the other four died within 6-10 days post CRRT treatment (figure 1A and 3A). The only surviving patient (P15) was admitted to the ICU because of critical kidney failure but not severe SARS-CoV-2-induced pneumonia, as mentioned above. The survival rate of patients who received CRRT treatment was only 6·3% (1/16), far lower than the ICU patients who did not receive CRRT treatment (31·1%, 14/45) (figure 3E, left panel). These data collectively suggested that CRRT might contribute to the high mortality of critical COVID-19 patients by inducing severe thrombocytopenia.

The progressive decrease in PLT in ICU patients suggested the possibility of HIT, since low-molecular-weight heparin (enoxaparine, LMWH) is routinely used as an anticoagulant in CRRT therapy in the hospital. LMWH is also widely used in elderly, bedridden ICU patients for the prevention of thrombosis and the management of coagulopathy and disseminated intravascular coagulation (DIC). In all 45 ICU patients without CRRT, 27 patients received LMWH. A higher but not statistically significant (*P*=0·09) percentage of these patients had severe thrombocytopenia (PLT≤50×10^2^/L) than did patients without LMWH (37·0%, 10/27 vs 11·1%, 2/18) (table 2). A lower survival rate was also observed in the non-CRRT patients exposed to heparin compared with non-CRRT patients without heparin exposure (25·9%, 7/27 vs 38·9%, 7/18) (figure 3E, right panel). Consistent with the fact that a smaller dose of LMWH was applied in non-ICU patients just for flushing of the central venous catheter, a mild PLT decrease was found only in four out of 93 non-ICU patients who received heparin, while the lowest number was 50-117×10^2^/L (figure S1B). A progressive PLT decrease to a level around 50×10^2^/L was observed in only one of these four patients. These findings indicate that heparin exposure may be a risk factor for severe COVID-19.

**Table 2.** Platelet counts and heparin-involved treatments.

| Treatment Group | Platelet Count ($\times 10^9/L$) | | | PLT$\leq 50 \times 10^9/L$<br>& Fall$>50\%$ |
| --- | --- | --- | --- | --- |
| | $\leq 50$ | $>50$ to $\leq 125$ | $>125$ | |
| CRRT ICU Patients (n=16) | 13 (81.3%) | 1 (6.3%) | 2 (12.5 %) | 9 (56.3%) |
| Non-CRRT ICU Patients (n=45) | 12 (26.7%) | 16 (35.6%) | 17 (37.8%) | 10 (22.2%) |
| I: Heparin (n=27) | 10 (37.0%) | 9 (33.3%) | 8 (29.6%) | 9 (33.3%) |
| II: w/o Heparin (n=18) | 2 (11.1%) | 7 (38.9%) | 9 (50.0%) | 1 (5.6%) |
| P | 0.08586 | 0.7578 | 0.2162 | 0.03433 |
| Non-ICU Patients (n=93) | 1 (1.1%) | 15 (16.1%) | 77 (82.8%) | 1 (1.1%) |
| I: Heparin (n=63) | 1 (1.6%) | 11 (17.5%) | 51 (81.0%) | 1 (1.6%) |
| II: w/o Heparin (n=30) | 0 (0%) | 4 (13.3%) | 26 (86.7%) | 0 (0) |
| P | N/A | 0.7668 | 0.5703 | N/A |
| P (Non-CRRT ICU vs Non-ICU) | 4.724e-07 | 0.03349 | 5.67e-07 | 5.334e-05 |
| P (Non-CRRT ICU I vs Non-ICU I) | 1.309e-05 | 0.1067 | 5.701e-06 | 5.309e-05 |
| P (Non-CRRT ICU II vs Non-ICU II) | 0.1356 | 0.07381 | 0.008523 | 0.375 |

### High risk of HIT in critical COVID-19 patients

Heparin exposure is correlated with the severe thrombocytopenia, suggesting that HIT might be responsible for severe thrombocytopenia in COVID-19. To substantiate this hypothesis, HIT antibodies against heparin-PF4 complexes, a marker of HIT, were analyzed using an ELISA kit. Higher levels of HIT antibodies were detected in the available sera from 14 COVID-19 ICU patients (sera from all ICU patients were always unavailable due to death or discharge), but not in mild COVID-19 patients, convalescent COVID-19 patients, or healthy people (figure 4A). To our surprise, an elevated HIT antibodies level was also observed in some of these ICU patients before CRRT implementation, or before heparin exposure in non-CRRT patients, suggesting that heparin pre-exposure was not absolutely required to induce HIT in critical COVID-19 patients (figure 4B). Accordingly, among 11 heparin-naïve nonsurvivors, seven patients experienced a progressive decrease in PLT, and four of them had a PLT decrease of more than 50% (figure 4C).

**Figure 4.**
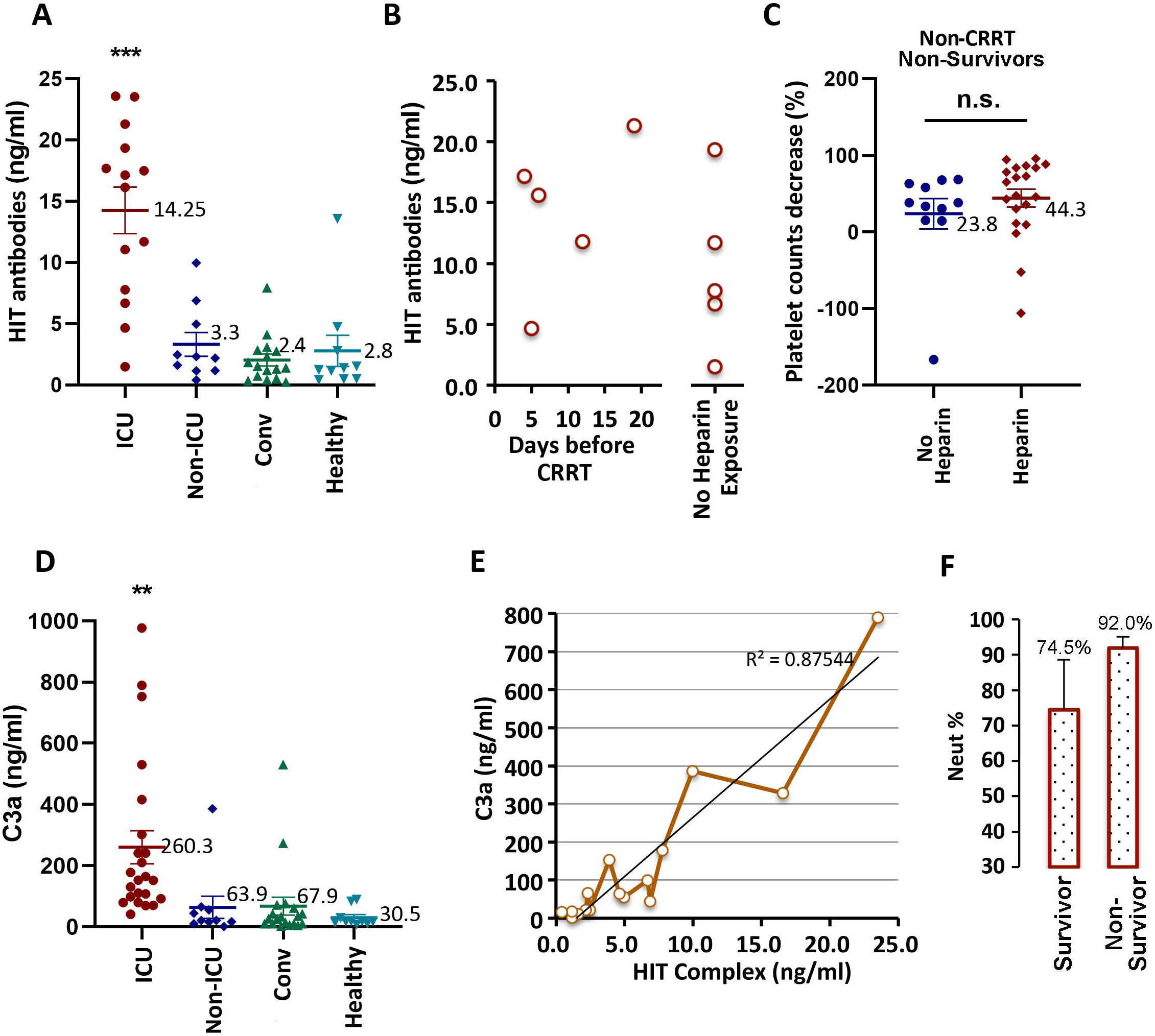
Heparin-induced thrombocytopenia in critical COVID-19 patients. (A) Level of heparin-PF4 complexes antibodies (HIT antibodies) in serum of critical patients (ICU), severe patients (non-ICU), convalescent patients (recovered non-ICU) and healthy controls. (B) HIT antibodies levels in patients before CRRT (CRRT patients) and nonsurviving patients without CRRT or other heparin exposure. (C) PLT decrease in nonsurviving COVID-19 patients without CRRT, with or without heparin exposure. (D) As in (A), serum C3 levels were determined. (E) The correlation between the level of HIT antibodies and C3a is fitted by a regression line with R^2^=0·87544. (F) Neutrophil percentage among white cells in survivors and nonsurvivors is shown. The data are presented as the mean ± SEM and were analyzed using a two-tailed Student’s *t-*test. Differences were considered significant at *\*\*P*<0·01, *\*\*\*P*<0·001, as indicated.

Consistent with the elevated HIT antibodies, complement activation shown by C3a accumulation was also observed in these patients (figure 4D). The concentration of serum HIT antibodies was largely correlated with C3a levels (figure 4E). It has been reported that the release of heparin from master cells could be induced by complement activation, which may indicate the potential role of endogenous heparin in spontaneous HIT after viral infection.^22,23^ Moreover, as reported by other studies, a higher percentage of neutrophils was observed in nonsurviving critical COVID-19 patients, which may further contribute to the severity of HIT, but not in surviving critical COVID-19 patients (figure 4F and S1D).^10,24^ In addition to laboratory tests, venous and arterial complications, including deep venous thromboembolic complications, were observed in patients who died of COVID-19 by autopsy.^25^ Among three nonsurviving ICU patients who received an autopsy, hyaline thrombus and other thrombi were found in patient P12, who showed a progressive decrease in PLT. Bleeding in lung tissues but no thrombi were observed in patient P17 with constantly low PLT counts (figure S1A). Furthermore, no obvious bleeding or thrombi occurred in patient P5, who had a significant decrease in PLT counts, but the lowest point was well above 100×10^2^/L. Confirmed vein thromboembolism (VTE) in ICU COVID-19 patients by CT pulmonary angiography (CTPA) or ultrasonography was also reported recently.^26^

Although the platelet activation assay was not performed because of the limited capacity of Huoshenshan Hospital, current results still strongly suggested that HIT or spontaneous HIT, induced by the virus or a secondary bacterial infection, may occur in COVID-19, which would be significantly boosted and aggravated by further heparin exposure with a high dose, thereby resulting in a fatal outcome.

## Discussion

Severe thrombocytopenia and thrombosis-induced systemic clotting in severe and critically ill COVID-19 patients resulted from complicated factors that greatly enhanced the risk of COVID-19. SARS-CoV-2 entry and infection may directly lead to endothelial damage, thereby triggering platelet activation and aggregation, a coagulopathy mechanism similar to that reported for SARS-CoV.^27^ Moreover, the serious complications of COVID-19 also result in elevated levels of D-dimer,^28^ a fibrin degradation product that indicates DIC, and antiphospholipid antibodies, an autoantibody that induces thrombi in large veins and arteries.^29^ All these factors contribute to increased platelet consumption and eventually thrombocytopenia.

Here, we show that HIT occurred frequently in severe COVID-19 patients who received heparin-involved therapy, or were heparin-naïve (as spontaneous HIT). The widely used diagnostic criteria of HIT include: 1) HIT antibodies to complexes of heparinoid (heparin or similar glycosaminoglycans) and PF4; 2) HIT antibodies-mediated platelet activation; 3) a progressive PLT decrease of at least 40-50% or more from baseline; 4) onset of thrombocytopenia occuring within five to ten days after initiation of heparin or 24 to 48 hours after heparin re-exposure; and 5) thrombosis in patients treated with heparin.^20,30,31^ In this study, severe thrombocytopenia (PLT<50×10^2^/L), a >50% of PLT decrease, rapid decrease upon CRRT treatment (high level of heparin exposure), high levels of HIT antibodies, and venous, arterial and even deep venous thrombi in patients who died of COVID-19 basically fits the clinical definition of HIT. Notably, HIT antibodies and a progressive decrease in PLT were also detected in heparin-naïve patients (i.e., before CRRT or other heparin exposure), even in non-ICU patients (figure 4B), indicating the occurrence of spontaneous HIT in COVID-19 patients, which probably results from virus itself or a secondary bacterial infection, such as PF4-conjugated *Staphylococcus aureus* or *Escherichia coli*, or severe tissue damage.^18^ Immune thrombocytopenic purpura (ITP), an autoimmune disorder similar to HIT, has been reported in HKU1 coronavirus infected patient.^32^ In non-ICU patients, a progressive PLT decrease is rarely occurred, either because of the less severe viral and secondary bacterial infection or the far lower level of heparin exposure, since heparin was primarily administered for flushing the central venous catheter but not for prevention blood clots and treatment of venous thrombosis in ICU patients. The moderate PLT decrease in non-ICU patients with HIT is transient and can be recovered during convalescence. Although a protective effect of heparin-involved anticoagulation in severe COVID-19 has been reported by Tang et al.,^33^ the potential risk of HIT should be cautioned when it’s used to treat critical ICU patients with a high dosage.

Our findings strongly indicated the occurrence of HIT in severe COVID-19, although current evidence did not fully meet the rigorous definition of spontaneous HIT proposed by Warkentin et al.^20^ because of the limited medical and laboratory capacity in *de novo* established Huoshenshan Hospital for COVID-19 patients. The platelet serotonin-release assay (SRA) for highly pathogenic HIT in sera containing both heparin-dependent and heparin-independent platelet activation antibodies, should be performed later, which will provide more solid evidence of HIT in COVID-19.^34^

In patients with HIT, anti-heparin-PF4 IgG could be generated as early as the first day of heparin treatment, indicating a preimmunization by antigens highly similar to heparin-PF4 complexes. This preimmunization may be resulted from spontaneous HIT. For preimmunized patients, subsequent exposure to a high dosage of heparin in anticoagulation therapy, such as CRRT and coagulation control, will significantly boost the preformed B cells via a lot of heparin-PF4 complexes, thereby prompting the activation and consumption of platelet, resulting in progressive thrombocytopenia and extensive coagulation in deteriorating COVID-19 patients. This may explain the high mortality in critically ill patients receiving CRRT (93·8%, 15/16). As an important supportive treatment for critically ill patients with sepsis or AKI, CRRT is used to efficiently remove proinflammatory mediators accumulated in the blood and maintain fluid balance in hemodynamically unstable patients,^35^ which has been recommended for renal failure and renal replacement therapy in the Chinese Clinical Guidance for COVID-19.^36^ However, accumulating clinical data showed that there were nearly no survivors among critical COVID-19 patients who received CRRT treatment, indicating that patients did not benefit from CRRT.^6,8,11^ Here, we show that serious HIT may be responsible for the high fatality of CRRT in critical COVID-19 patients. Therefore, we suggeste that the clinical occurrence of HIT in COVID-19 patients, especially critical patients, should be detected and monitored carefully. For those COVID-19 patients strongly suspected of having HIT, the physician should stop all heparin-involved therapy and initiate an alternative anticoagulant other than heparin, such as platelet aggregation inhibitor prostacyclin, direct thrombin inhibitors (e.g., lepirudin), nonheparin glycosaminoglycan with anti-factor Xa activity (e.g., fondaparinux), or regional citrate anticoagulation, to avoid the risk of HIT-induced thrombosis in COVID-19 patients.

### Patient consent

The informed consent and the approval from ethic committee of Huoshensan Hospital were obtained before the collection of the blood samples from COVID-19 patients.

## Data Availability

All data is available in the main text or the supplementary materials.

## Acknowledgments

The authors wish to thank the staff of Huoshenshan Hospital (Wuhan, China) for their unimaginable effort and devotion in COVID-19 pandemic and generous help in medical cases sharing and consultation.

## Author contributions

QM, YX and ZW collected COVID-19 patients clinical data. ZZ, HL and KL performed the ELISA detection. XZ, GW, and YH analyzed the laboratory test results. WC, QD and SZ provided laboratory detection materials, methods and equipment. CC, QM, SZ, and LY interpreted clinical therapeutic outcome. XB was consulted on COVID-19 autopsy. CC and TG made the figures. XL and CC wrote the manuscript.

## Supplementary Materials

### Supplementary figure and table legends

**Figure S1.**
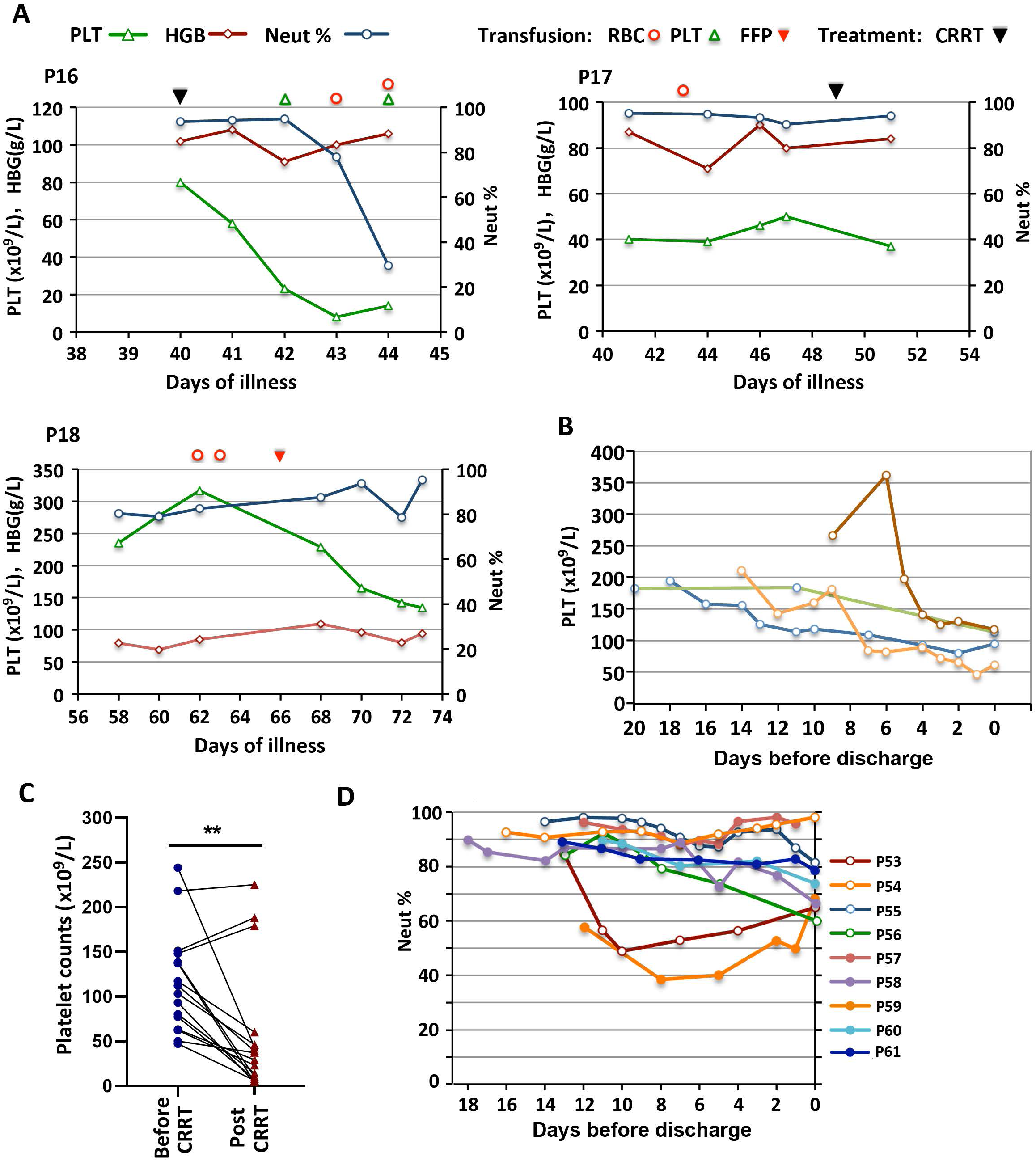
Platelet counts and neutrophil percentages in the COVID-19 patients. (A) Day-by-day platelet count, hemoglobin level, and neutrophils percentage in three ICU nonsurvivors (P16-P18). (B) Day-by-day platelet count in 4 non-ICU patients with moderate progressive platelet decrease. (C) The paired comparisons of platelet count before and after CRRT in all 16 critical COVID-19 patients with CRRT treatment. (D) Day-by-day neutrophil percentage among white cells in other surviving critical patients without CRRT treatment (P53-P61, corresponding to figure 2). Differences were considered significant at *\*\*P*<0·01, as indicated.

**Table S1.** Demographic and heparin-involved treatment of patients on admission.

| Treatment |  | Patient Number |  |  | Age |  |
| --- | --- | --- | --- | --- | --- | --- |
|  |  | Total | Male | Female | Average | SD |
| Critical Patients In ICU | Total | 61 | 41 | 20 | 72.41 | 10.4 |
|  | CRRT | 16 | 11 | 5 | 70.8 | 11.9 |
|  | Non-CRRT | 45 | 30 | 15 | 73 | 9.9 |
|  | Heparin | 27 | 18 | 9 | 72.3 | 7.4 |
|  | w/o Heparin | 18 | 12 | 6 | 74.2 | 14.9 |
| Non-ICU Severe Patients | Total | 93 | 53 | 40 | 70.1 | 11.01 |
|  | Heparin | 63 | 33 | 30 | 71.46 | 10.97 |
|  | w/o Heparin | 30 | 20 | 10 | 67.23 | 10.7 |

